# A polygenic predictor of baseline QTc is associated with sotalol-induced QT prolongation

**DOI:** 10.1101/2024.05.01.24306714

**Authors:** Megan C. Lancaster, Giovanni Davogustto, Edi Prifti, Claire Perret, Christian Funck-Brentano, Dan M. Roden, Joe-Elie Salem

## Abstract

**Background:** Drug-induced QT interval prolongation raises the risk of fatal arrhythmia and is a central issue in safety pharmacology. Yet individual QT drug response is highly variable, and risk stratification remains a challenge. We hypothesized that genetic factors underlying the baseline QT influence this variation in QT drug response.

**Methods:** A cohort of 990 healthy subjects was prospectively challenged with a single 80 mg oral sotalol dose. Exclusion criteria included abnormal ECG, cardiac disease, and QT-prolonging drug use. ECGs were obtained at baseline and 3 hours post-dose. QT response was defined as absolute change in Fridericia heart-rate corrected QT (ΔQTc). Plasma sotalol was measured 3 hours post-dose. Subjects were genotyped on the MEGA array. Principal component analysis of genotypes was used to define genetic ancestry. A polygenic risk score (PRS) for the baseline QTc comprised of 465,399 common variants was calculated from assayed and imputed genotypes. The difference in PRS between subjects with high QT response (ΔQTc≥60 ms) and those with ΔQTc<60ms was assessed by Mann Whitney test. A multivariable regression model was used to assess the association between PRS and ΔQTc after covariate adjustment. *Results:* Of the 990 subjects, 978 (99%) passed genomic quality control and were included in analysis. 62% were female (n=607), with median age 23 (interquartile range (IQR) 21-33). The median ΔQTc 3 hours post-sotalol was 21 ms (IQR 12-30). Ten subjects (1%) had ΔQTc≥60 ms, and their PRS was significantly higher compared to those with ΔQTc<60 ms (P=0.0082). In the regression model, PRS was positively associated with ΔQTc (P=0.0002).

**Conclusion:** Common genetic variants associated with the baseline QT also capture part of the repolarization response to sotalol. This adds to the emerging concept that the genetic architecture of a trait can predict drug response.

---

Long QT intervals predispose to the morphologically distinctive ventricular tachycardia, Torsade de Pointes (TdP).^1^ Exaggerated QT prolongation can occur after exposure to numerous commonly-used drugs, most of which result in block of the rapid delayed rectifier (I_Kr_) channel.^1^ Following drug exposure, an increase in the QT corrected for heart rate (QTc) to >500 msec or of >60 msec versus baseline has been used as a marker for TdP risk.^1^ However, interindividual QT response to a drug is highly variable, and risk stratification of low-versus high-risk for extreme QT prolongation remains a challenge. Large genome-wide studies have identified sets of single nucleotide variants (SNV) influencing baseline QTc, and polygenic risk scores (PRS) integrating their effects have been developed to capture this influence.^2^ Here, in a prospective European ancestry cohort of almost 1000 subjects, we examine the relationship between a QTc-PRS (PRS for QTc prior to drug) and QTc prolongation in response to a standardized challenge with the I_Kr_-blocking drug sotalol.

Using the GENEREPOL^3^ cohort (NCT00773201) of 990 healthy individuals with a normal baseline ECG who were prospectively challenged with a single oral dose of 80mg sotalol, we measured the QTc change between baseline and 3 hours post-dosing (ΔQTc; Fridericia correction) as previously described.^3^ The plasma concentrations of sotalol 3 hours post-dosing and of potassium were also measured. Subjects were genotyped using the MEGA^ex^ array, with 978 samples (99%) passing standard quality control and 12 excluded for the following: heterozygosity F>0.19 (n=1), patient duplication (n=1), missing sample data (n=1), and unconfirmed genetic/clinical record sex match (n=9). We constructed a QTc-PRS consisting of the 465,399 SNV from a previously generated 1,110,495 SNV QTc-PRS^2^ that were present in our imputed genomes at major allele frequency >0.01 and with imputation r^2^>0.7. We generated a multivariable regression model (ANCOVA, best model approach selected by Bayesian Schwarz information criterion) studying the association of ΔQTc with QTc-PRS, while adjusting for sex, age, baseline QTc, plasma potassium, plasma sotalol concentration, and the first ten principal components of genetic ancestry as independent variables. P-value<0.05 was considered significant.

The analyses included 978 healthy subjects (607, 62% female), with median age 23.6 years (interquartile range (IQR) 21.0-32.7). Median baseline QTc was 388 msec (IQR 376-399). Three hours post-sotalol intake, the median ΔQTc was 20.7 msec (IQR 12.0-30.3). Ten subjects (1.02%) had ΔQTc≥60msec (“extreme responders”). None had a post-sotalol QTc above 500 msec. In the extreme responder group, the QTc-PRS was significantly greater than in those with a smaller drug response, or “normal responders” (mean z-score 0.75 vs. −0.0077, *p*=0.008, Mann Whitney test, Figure 1A-B). We found that QTc-PRS was able to discriminate normal responders vs. extreme-responders with an area under the receiver operating curve of 0.74 (95% Confidence Interval (CI) 0.62-0.87). In this cohort, a QTc-PRS threshold of −0.4 had a 100% (CI 100-100%) sensitivity and 100% negative predictive value (CI 100-100%) to detect extreme responders, leaving 32.1% (n=314) below this threshold (Figure 1C for the detailed diagnostic properties of the main QTc-PRS thresholds). Furthermore, in the multivariable regression model (adjusted r^2^=0.35, p<0.0001), ΔQTc was associated with QTc-PRS (β_standardized-coefficient_=0.095±0.026, *p*=0.0002) while including the significant contribution of sex (β=-0.42±0.026 for male vs. female, *p<*0.0001), age (β=-0.080±0.026, *p*=0.002), plasma sotalol level (0.36±0.026, *p*<0.0001), and plasma potassium (0.082±0.026, *p*=0.002). In this latter model, the baseline QTc and the first ten principal components of genetic ancestry were not significant.

**Figure 1:**
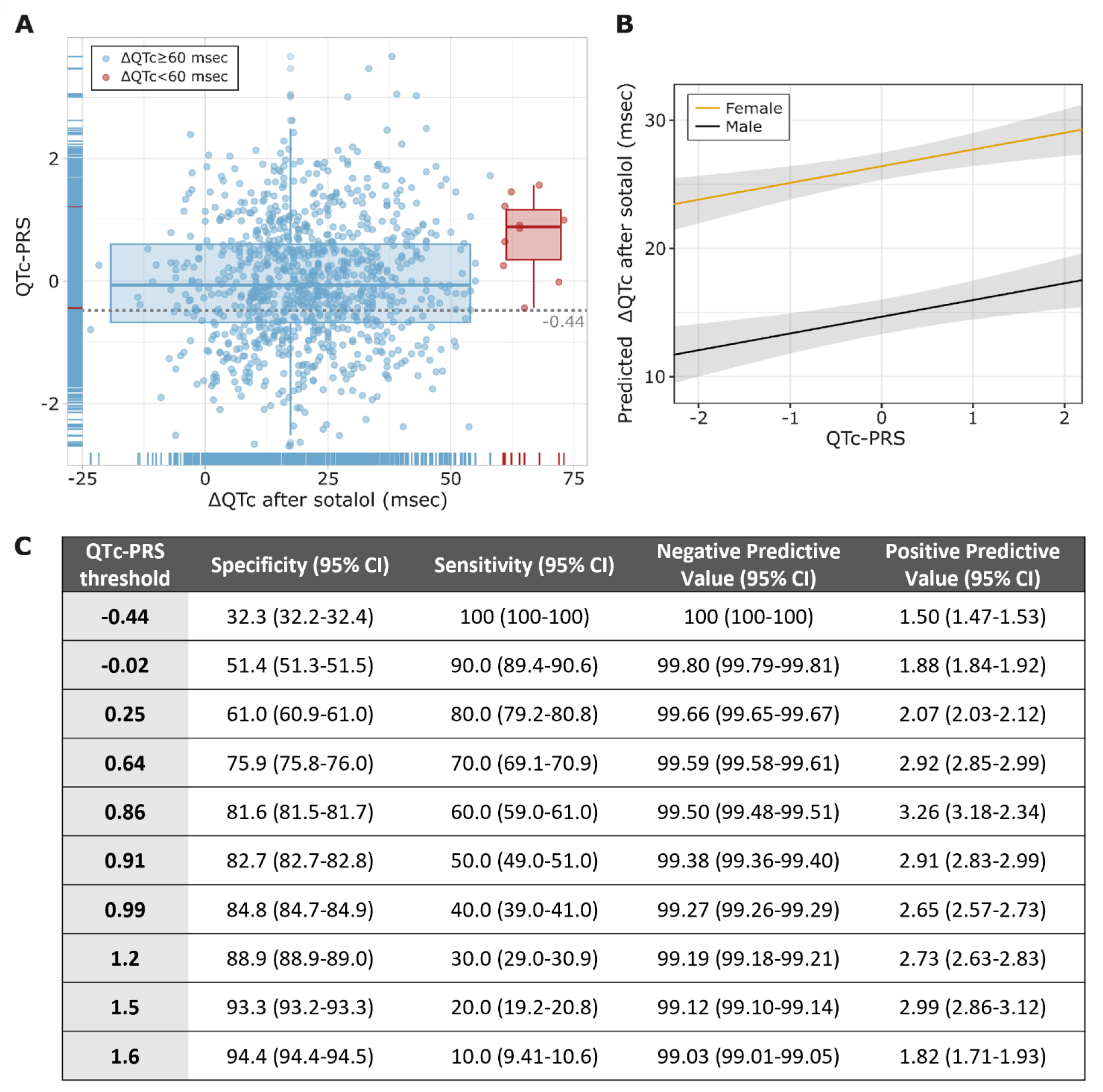
**(A)** Interindividual variation in the QTc response to 80 mg of sotalol (ΔQTc) versus the polygenic risk score for the baseline QTc (QTc-PRS, N=978). The QTc-PRS is higher in extreme-responders (ΔQTc ≥60 msec, red, n=10) than in normal responders (blue, n=968) by the two-sided Mann-Whitney test (*P*=0.0082) **(B)** The predicted ΔQTc (msec) as a function of the QTc-PRS (z-score) in females and males in multivariable regression model including age, plasma sotalol concentration, plasma potassium concentration, baseline QTc, and the first 10 principal components of genetic ancestry (*p*_model_ <0.0001 and *p*_qt-PRS_=0.0002). Gray indicates 95% confidence intervals. **(C)** Specificity, sensitivity, negative and positive predictive values (PPV) in identifying extreme-responders using various QTc-PRS thresholds. PRS = polygenic risk score, QTc-PRS = polygenic risk score for the baseline QT interval, corrected for heart rate (QTc).

Our results show that the common genetic variants associated with QTc prior to drug exposure also capture part of the repolarization response (ΔQTc) dynamics following exposure to an I_Kr_ blocking-drug in a cohort of approximatively a thousand participants, with ECG acquisition and QTc measurement quality standards similar to those applied in thorough QT-studies, which are usually performed on approximately 20-40 patients. This finding is concordant with a previous report that a simplified QTc-PRS (61 SNV) was associated with the extent of QT-prolongation after drug challenge in 22 healthy subjects, and was also associated with TdP risk in 216 TdP cases and 771 drug-exposed controls.^4^ The idea that a baseline PRS predicts drug response has also been reported for a baseline PRS for coronary artery disease and outcomes in PCSK9 inhibitor therapy, and a baseline PRS for depression and response to antidepressants.^5^ Thus, the data we report not only define how QTc-PRS predicts ΔQTc but also add to the emerging concept that the genetic architecture of common disease or traits can be used to predict drug response. Additionally, the potential of a very high QTc-PRS negative predictive value to rule-out extreme responders in this proof-of-concept work is promising and deserves further validation, e.g., to confirm that QTc-PRS could be used to avoid QTc monitoring in a predicted low risk population.

## Data Availability

Summary level data produced in the present study are available upon reasonable request to the authors. To avoid subject reidentification, subject-level data will not be available.

## Notes

### Competing Interest Statement

The authors have declared no competing interest.

### Clinical Trial

NCT00773201

### Funding Statement

The clinical study was supported by a
grant from INSERM/DHOS (Clinical and
Translational Research award 2007, RCT07002)
and by the French Ministry of Health (PHRC2008-AOR08004). Dr. Lancaster was supported by the National Institutes of Health T32 HG008962. Dr. Davogustto was supported by the American Heart Association SFRN grant.

### Author Declarations

The study protocol and all methods applied were approved by the Committee for the Protection of Human Subjects of Paris Ile de France V (Paris, France) and prior written informed consent was obtained from all subjects after being fully informed regarding the nature and risks of the study. We did the study in accordance with the principles of the international conference on harmonization guidelines on good clinical practice and the world medical association declaration of Helsinki.

